# Does WASH FIT improve water, sanitation, and hygiene and related health impacts in healthcare facilities? A systematic review

**DOI:** 10.1101/2024.04.05.24305396

**Authors:** Hannah Lineberger, Ryan Cronk, Sena Kpodzro, Aaron Salzberg, Darcy M. Anderson

## Abstract

**Introduction:** Environmental health services (e.g., water, sanitation, hygiene, cleaning, waste management) in healthcare facilities are important to improve health outcomes and strengthen health systems, but coverage gaps remain. The World Health Organization and United Nations Children’s Fund developed WASH FIT, a quality improvement tool, to help assess and improve environmental health services. Fifty-three countries have adopted it. However, there is little evidence of its effectiveness. This systematic review evaluates whether WASH FIT improves environmental health services or associated health outcomes and impacts.

**Methods:** We conducted database searches to identify relevant studies and extracted data on study design, healthcare facility characteristics, and inputs, activities, outputs, outcomes, and impacts associated with WASH FIT. We summarized the findings using a logic model framework and narrative synthesis.

**Results:** We included 31 studies in the review. Most inputs and activities were described qualitatively. Twenty-three studies reported quantitative outputs, primary WASH FIT indicator scores, and personnel trained on WASH FIT. Nine studies reported longitudinal data demonstrating changes in these outputs throughout WASH FIT implementation. Six studies reported quantitative outcomes measurements; the remainder described outcomes qualitatively or not at all. Common outcomes included allocated funding for environmental health services, community engagement, and government collaboration, changes in knowledge, attitudes, or practices among healthcare staff, patients, or community members, and policy changes. No studies directly measured impacts or evaluated WASH FIT against a rigorous control group.

**Conclusions:** Available evidence is insufficient to evaluate WASH FIT’s effects on outputs, outcomes, and impacts. Further effort is needed to comprehensively identify the inputs and activities required to implement WASH FIT and to draw specific links between changes in outputs, outcomes, and impacts. Short-term opportunities exist to improve evidence by more comprehensive reporting of WASH FIT assessments and exploiting data on health impacts within health management information systems. In the long term, we recommend experimental studies. This evidence is important to ensure that funding invested for WASH FIT implementation is used cost-effectively and that opportunities to adapt and refine WASH FIT are fully realized as it continues to grow in use and influence.

**Highlights:**

- WASH FIT is highly influential, but little is known about its effectiveness
- We reviewed WASH FIT’s effects on environmental health service outputs and health impacts
- Nine studies measured outputs longitudinally; none directly measured health impacts
- No studies compared WASH FIT’s performance against a rigorous control group
- Evidence is insufficient to assess WASH FIT’s effects on outputs or health impacts

**Graphical abstract:** 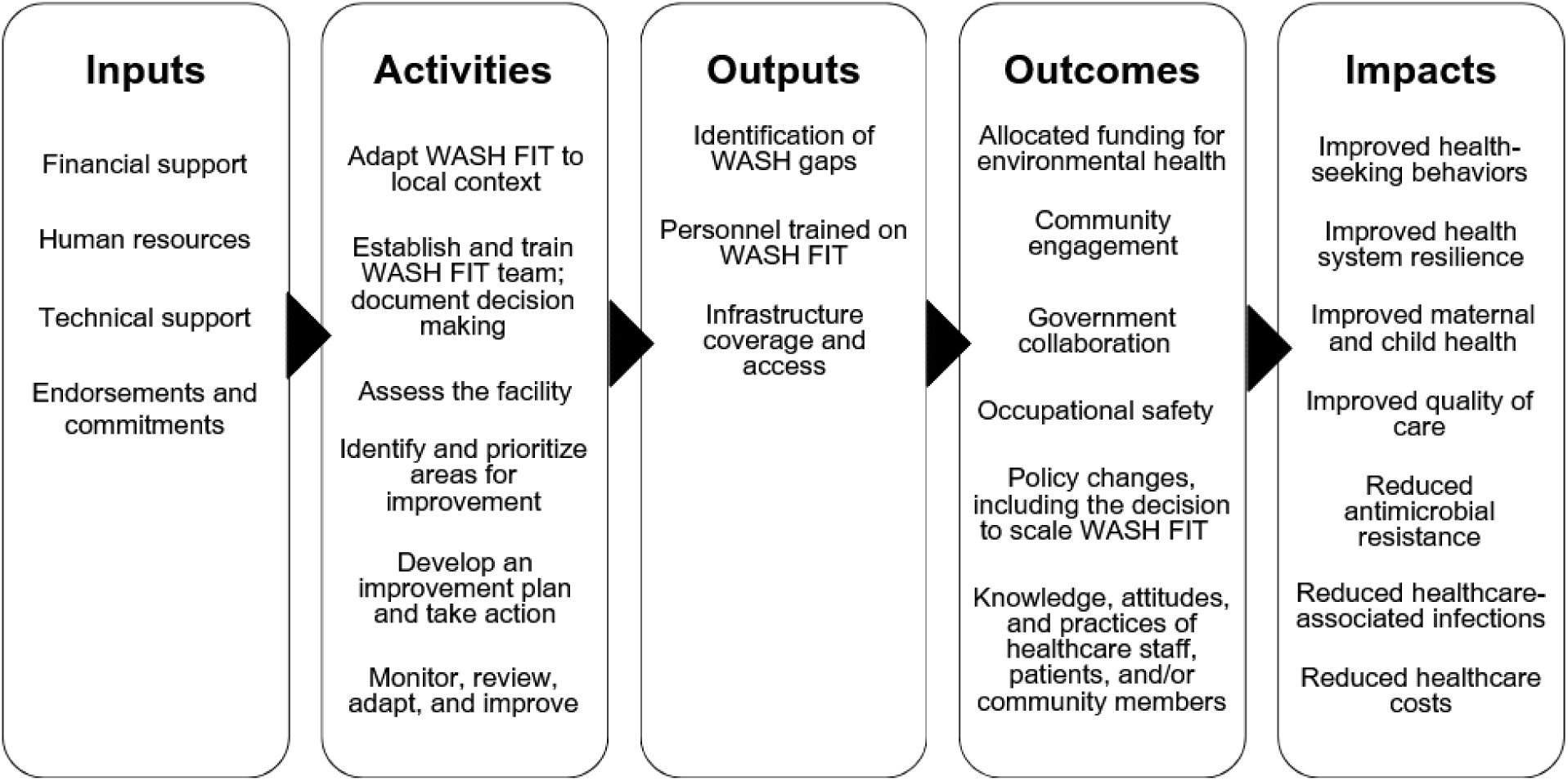

## 1. Introduction

Environmental health services, including water, sanitation, hygiene, cleaning, and waste management, are critical for safe healthcare facilities. They reduce the risk of healthcare-associated infections, which can help reduce antimicrobial resistance (AMR) and healthcare costs (Hutton et al., 2024; Watson et al., 2019). Environmental health services are associated with improved quality of care, care seeking, and patient and healthcare worker satisfaction (Anderson et al., 2023; Bouzid et al., 2018; Fejfar et al., 2021). Investing in environmental health services in healthcare facilities is recommended as an important intervention to improve maternal and child health and health system resilience (Velleman et al., 2014).

Despite the benefits of environmental health services, gaps in coverage remain. Twenty-two percent of healthcare facilities lack basic water services, 49% lack basic hygiene services, and 10% have no sanitation services (WHO/UNICEF, 2023). Healthcare facilities in low- and middle-income countries experience the largest deficiencies and disproportionate infectious disease burdens, contributing to a cycle of poverty and inequitable health outcomes (Bhutta et al., 2014; WHO/UNICEF, 2023).

To make progress toward universal access, the World Health Organization (WHO) and the United Nations Children’s Fund (UNICEF) developed the Water and Sanitation for Health Facility Improvement Tool (WASH FIT) in 2017, with a second edition released in 2022. WASH FIT is a continuous quality improvement tool to help evaluate and improve environmental health services in healthcare facilities in resource-limited settings (WHO/UNICEF, 2022a). WASH FIT implementation follows a five-step cycle: 1) establish and train the team; document decisions; 2) assess the facility; 3) identify and prioritize areas for improvement; 4) develop an improvement plan and act; and 5) monitor, review, adapt, and improve. This cycle is intended to be iterated every 6-12 months for continuous quality improvement.

Integrated into WASH FIT is an assessment tool containing indicators to assess coverage and access to environmental health services. This tool is used in Step 2 of the WASH FIT cycle. These indicators guide improvement plans (Step 3) and are intended to be monitored and reassessed (Step 5) to evaluate progress as part of continuous quality improvement. The WASH FIT first edition assessment tool comprises 66 indicators across four core domains: water, sanitation (divided into Part A for “sanitation” and Part B for “healthcare waste”), hygiene (divided into Part A for “hand hygiene” and Part B for facility environment, cleanliness, and disinfection), and management (WHO/UNICEF, 2017). The second edition comprises 95 indicators across five primary domains (water, sanitation, healthcare waste management, environmental cleaning), two domains needed to support WASH infrastructure and practice (energy and the environment; management and workforce), and two cross-cutting themes (climate resilience and equity and inclusiveness) (WHO/UNICEF, 2022a).

WASH FIT has been implemented in 53 countries as of 2024 and is one of the most widely adopted and influential tools for prioritizing investments in environmental health services in healthcare facilities. Individual country programs have received investments of millions of dollars (Terre des hommes, 2018; USAID, 2023); the total amount of health financing influenced by WASH FIT globally is difficult to quantify but likely totals billions of dollars.

Yet little is known about its effectiveness in changing target WASH service-level outputs, health impacts, or the causal pathways by which those changes may occur. Most research on WASH FIT describes its use as a one-time assessment tool to identify service gaps and prioritize resources (Ashinyo et al., 2021; Doku et al., 2022; Hirai et al., 2021). Fewer studies have evaluated its potential outcomes and impacts or described the inputs and activities needed to achieve those long-term results (Aung & Chettry, 2021; Kabir et al., 2023; Weber et al., 2018). There are currently two proposed conceptual frameworks for WASH FIT impact pathways, but neither has been empirically vetted (Weber et al., 2019; WHO/UNICEF, 2022a).

We conducted a systematic review to evaluate whether WASH FIT improves service-level outputs for environmental health services or health outcomes and impacts. Our objectives are to (1) describe the study designs and WASH FIT domains measured in WASH FIT studies; (2) document the inputs and activities required to deliver WASH FIT; and (3) evaluate the effect of WASH FIT on outputs, outcomes, and impacts.

## 2. Material and methods

### 2.1. Study overview

We systematically reviewed peer-reviewed and grey literature on WASH FIT implementation and evaluation. We extracted data on WASH FIT study types and locations, length of WASH FIT implementation, and WASH FIT domains measured in each study to assess study designs and WASH FIT domains. To determine the inputs and activities required to deliver WASH FIT, we extracted data on WASH FIT inputs and activities. We mapped activities to the WASH FIT’s intended five-step continuous quality improvement cycle (WHO/UNICEF, 2022a) and inductively developed categories for inputs based on observed similarities in the data. We used a similar approach to compile and evaluate WASH FIT outputs, outcomes, and impacts.

### 2.2. Conceptual framework

We used a logic model to guide our data extraction and narrative analysis. Logic models visually depict the relationships between an intervention’s inputs, activities, outputs, outcomes, and impacts (Figure 1). They help identify and communicate causal pathways related to a program or policy and are frequently used for program monitoring and evaluation (W.K. Kellogg Foundation, 2004.) We used a logic model to document the inputs and activities required to deliver WASH FIT. We assessed outputs, outcomes, and impacts within the logic model framework to evaluate whether WASH FIT improved health impacts or service-level WASH outcomes.

**Figure 1.**
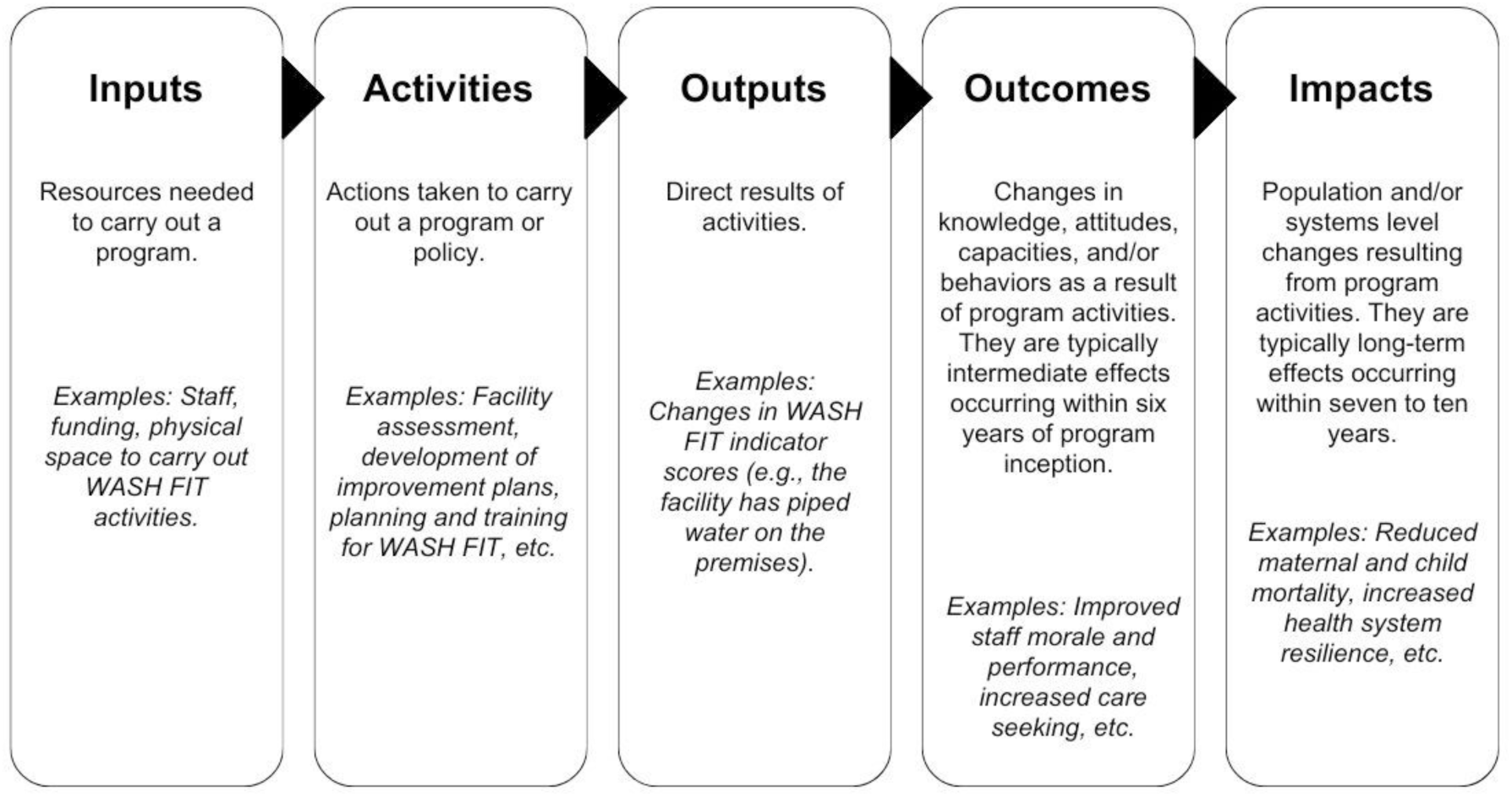
Defining Components of the logic model. Adapted from W.K. Kellogg Foundation (2004)

### 2.3. Search strategy

We conducted systematic searches of peer-reviewed and grey literature. We performed database searches in PubMed and Scopus for peer-reviewed literature on February 13, 2024. We searched by title and abstract in PubMed and title, abstract, and keyword in Scopus for four terms: “WASH FIT,” “WASH FAST,” “Water and sanitation for health facility improvement tool,” and “Water and sanitation for facility improvement tool.” We restricted our search to studies published in 2017 or later (the publication year for the first edition of WASH FIT).

For grey literature, we searched washinhcf.org, the largest online repository of resources related to environmental health services in healthcare facilities, curated by the WHO and UNICEF. Resources are tagged with keywords describing their content when uploaded. We searched washinhcf.org on February 6, 2024, for all resources tagged as WASH FIT. We identified additional references through a 2022 report on WASH FIT case studies (WHO/UNICEF, 2022b) and consultation with experts from the WHO and UNICEF.

### 2.4. Inclusion and exclusion criteria

We included studies that reported WASH FIT implementation in a healthcare facility in a low- or middle-income country and described at least one domain related to WASH FIT in the logic model (i.e., inputs, activities, outputs, outcomes, or impacts). Studies of all designs that used either the first or second edition of WASH FIT were eligible for inclusion. We restricted studies to English, French, and Spanish. We included only studies that reported implementing at least one step of the WASH FIT cycle. For example, we included some studies that partially implemented the WASH FIT cycle but did not report all five steps. However, we excluded studies that reported solely on pre-implementation processes (e.g., national-level training of trainers) without describing WASH FIT implementation in a healthcare facility. A single author screened all articles.

### 2.5. Data extraction and synthesis

Before data extraction, we developed, piloted, and revised the extraction form. The authors met to discuss the study goals and objectives and developed a preliminary form in Microsoft Excel. Two authors then piloted the form on five studies, met to discuss the challenges and shortcomings of the extraction form, and revised it accordingly. The final extraction included the following categories: study design and context (e.g., location, design, methods, healthcare facility characteristics), WASH FIT domains used in the assessment step, WASH FIT inputs and activities, and WASH FIT outputs, outcomes, and impacts. A single author extracted data from each study. The complete data extraction form is provided in Supplemental Information File 1.

We used narrative synthesis to address our three objectives. To describe the study designs and WASH FIT domains, we described similarities and differences in how each study measured and reported WASH FIT domains. To document inputs and activities, we organized extracted data in the logic model framework and inductively developed categories for inputs based on observed similarities in the extracted data. We mapped activities to the WASH FIT cycle (WHO/UNICEF, 2022a). There were insufficient data for a meta-analysis, so we organized extracted data on WASH FIT outputs, outcomes, and impacts in the logic model framework and inductively developed categories for outputs and outcomes based on observed similarities in the extracted data. We summarized the most common effects hypothesized in our logic model.

## 3. Results

### 3.1. Included studies

Our search yielded 156 unique articles. We identified 17 studies that were not identical but reported data on the same WASH FIT program (typically a grey literature report subsequently published in peer-reviewed literature). We included multiple studies on the same WASH FIT program only when they reported different data across inputs, activities, outputs, outcomes, and/or impacts. Where articles presented the same data, we excluded the older or less comprehensive version. Details of unique articles excluded based on duplicate data are provided in Supplemental Information File 1.

In total, 24 articles met our inclusion criteria. One article was a WHO report containing eight eligible case studies, each summarizing WASH FIT implementation in a different country (WHO/UNICEF, 2022b). We extracted and reported these case studies separately in our results for 31 studies included in our synthesis.

**Figure 2.**
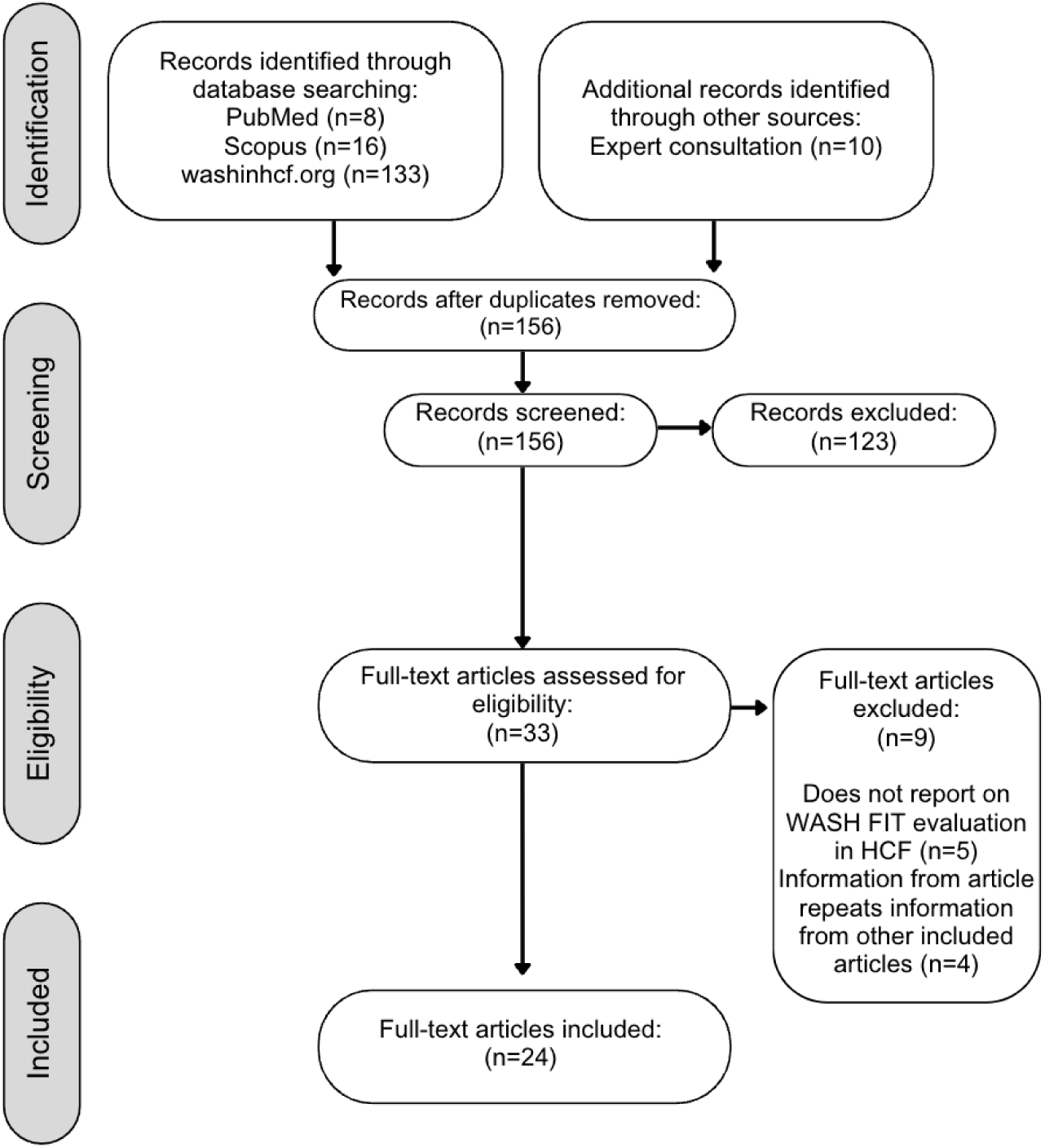
Flow chart of studies included in a review of WASH FIT effectiveness.

We found published reports on WASH FIT implementation and evaluation in 20 of the 53 countries where WASH FIT has been implemented as of 2024 (Figure 3). Studies from the African Region and South-East Asian Region represented approximately 71% (n=22) of the studies.

**Figure 3.**
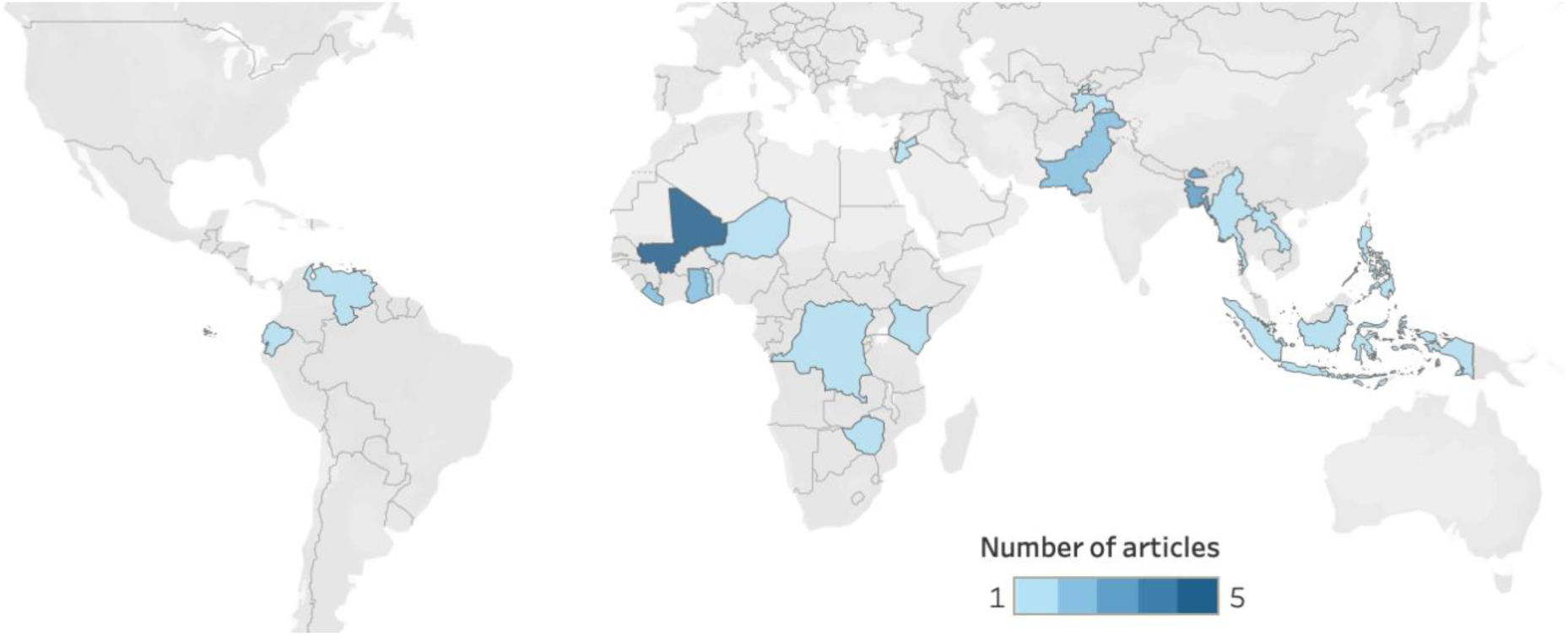
Geographic distribution of WASH FIT studies. Cropped regions of the map did not have any countries with WASH FIT studies.

More than half of the included studies (n=17, 55%) reported implementing WASH FIT in ten or more healthcare facilities, but sample sizes varied from one to 256. Of studies that reported healthcare facilities characteristics, WASH FIT implementation spanned all healthcare facility types (i.e., primary, secondary, tertiary), both publicly and privately ownership, and diverse types of health service provision, including inpatient, outpatient, maternity, neonatal, pediatric, medical, surgical, family planning, and laboratory services. The reporting of healthcare facility size varied. Some studies reported number of beds (n=4, 13%), some reported number of patient consultations (n=3, 10%), and others reported both (n=2, 6%). Fourteen studies (45%) reported no healthcare facility characteristics besides facility type or location. Of studies that reported location (n=14, 45%), 11 included HCFs in rural areas.

We identified ten studies of WASH FIT implementation in special settings or situations: three in refugee camps (Kabir et al., 2023; Morshed, 2019; WHO/UNICEF, 2022b) and seven as part of pandemic response or recovery for COVID-19 (Ashinyo et al., 2021; Hirai et al., 2021; Saadeh et al., 2022; WHO/UNICEF, 2022b), Ebola (Kanagasabai et al., 2021), or cholera (Ndumbi et al., 2020).

### 3.2. Study designs and follow-up time

Study designs comprised case study (n=17), cross-sectional (n=8), and quasi-experimental (n=6). Not all studies reported the length of WASH FIT implementation. Of those that did (n=19, 61%), WASH FIT implementation ranged from two days to three years, and most of these (n=11) reported implementing WASH FIT for one year or less. Studies of shorter durations typically used WASH FIT as a one-time assessment tool to identify gaps in environmental health service provision, set priorities, and inform the development of improvement plans. Nine studies (29%) recorded multiple rounds of WASH FIT assessment and systematically compared scores over time. Twelve studies (39%) completed one WASH FIT assessment to assess pre-improvement scores. The remaining ten studies (32%) did not specify how many assessments were conducted.

### 3.3. Evaluation of WASH FIT domains and indicators

In all studies, WASH FIT implementation and data collection was carried out before 2022 (the release date for the second edition), so we determined that all studies used the first edition. The aggregate “hygiene” and “sanitation” domains complicated our efforts to extract information on the specific WASH FIT domains measured and reported by each study. Fourteen studies reported disaggregated indicators for “hand hygiene” versus “facility environment, cleaning, and disinfection.” In contrast, two studies generically reported that they measured “hygiene” but did not specifically describe whether they measured “hand hygiene” indicators, “facility environment” indicators, or both. Similarly, 15 studies reported disaggregated indicators for “sanitation.” In contrast, three studies generically reported that they measured “sanitation” but did not specify whether they measured only “sanitation” or both “sanitation” and “healthcare waste” (Figure 4).

**Figure 4.**
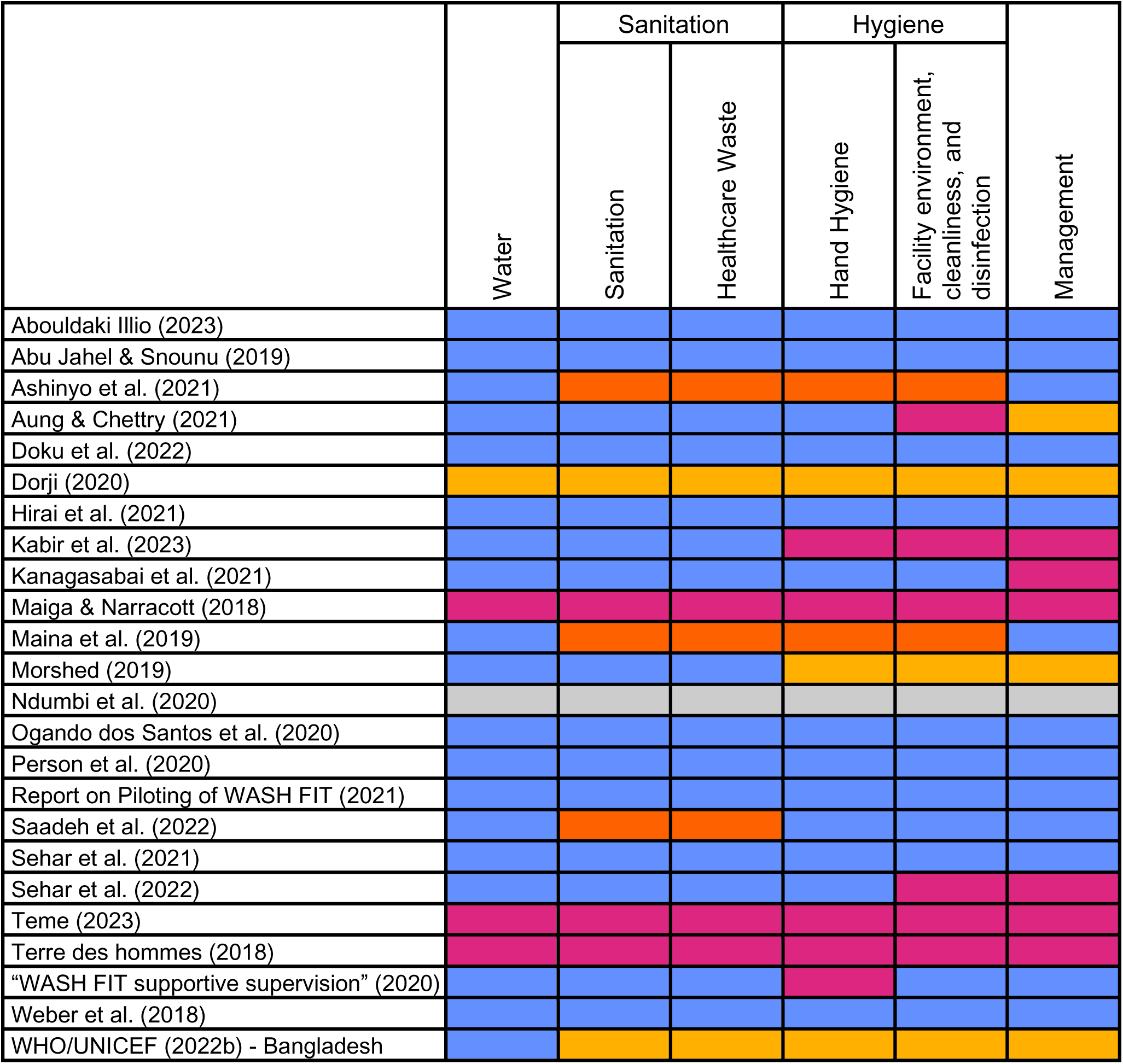
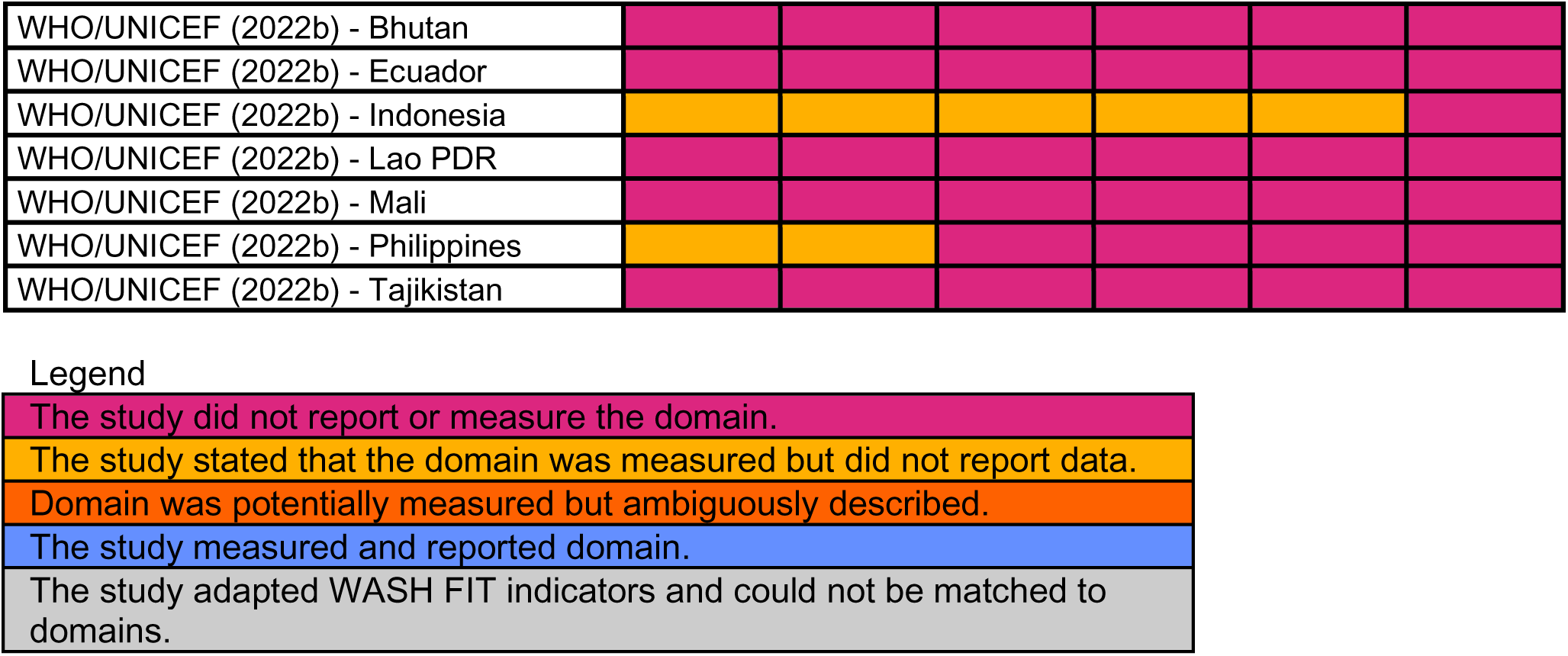
WASH FIT domains assessed in WASH FIT step 1 (assess the facility).

Nine articles reported on all WASH FIT first edition domains and sub-domains. Studies that did not report all domains were most likely to exclude the management or hand hygiene sub-domain. Not all studies explicitly reported the total number of WASH FIT indicators measured. Among those that did (n=16, 52%), the number of indicators ranged from seven to 67. Eight studies (26%) adapted WASH FIT indicators or supplemented them with indicators from other sources. For example, Ndumbi et al. (2020) reported using WASH FIT, but the indicators did not directly match any of the first edition WASH FIT indicators. Saadeh et al. (2022) measured 150 indicators, including WASH FIT and other indicators for infection prevention and control, COVID-19 safety, and pharmacy safety.

### 3.4. Inputs and activities required to deliver WASH FIT

Approximately 84% (n=26) of studies reported inputs and 100% (n=31) reported activities. Among these, most reported inputs and activities qualitatively. Case studies were most likely to report inputs, while cross-sectional studies were least likely to report them (Figure 5).

**Figure 5:**
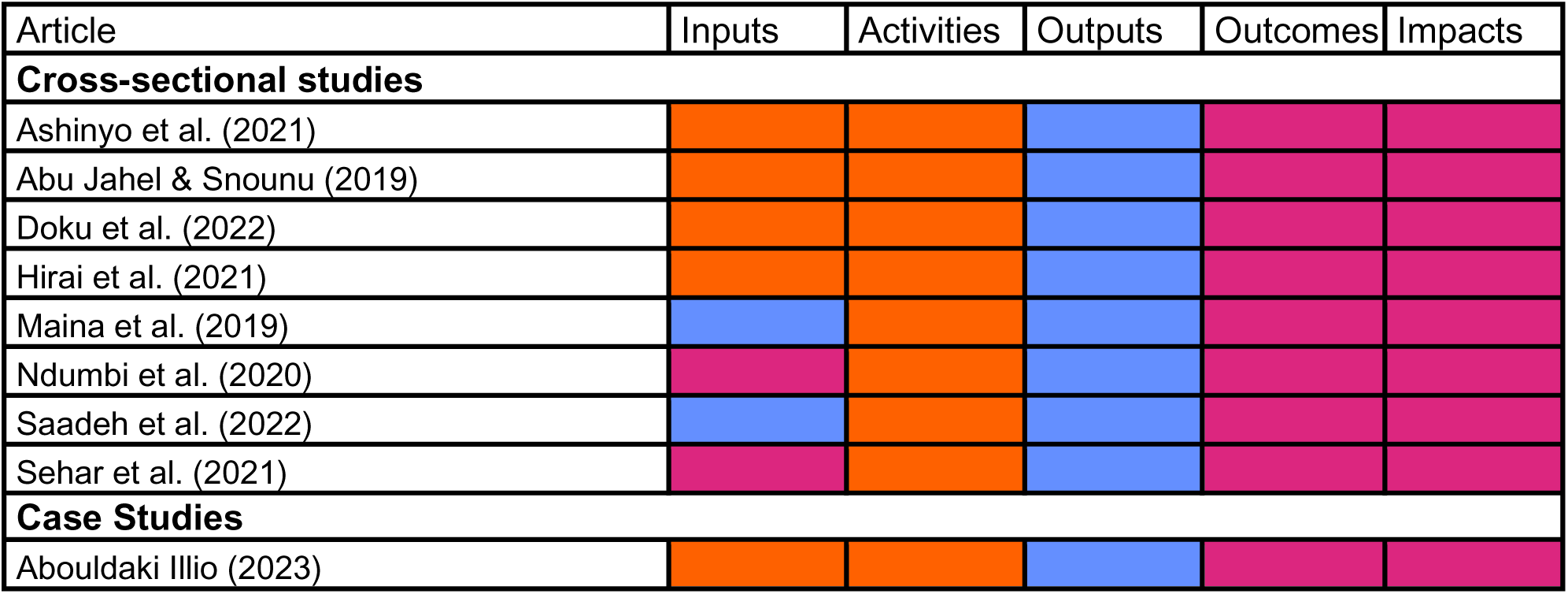
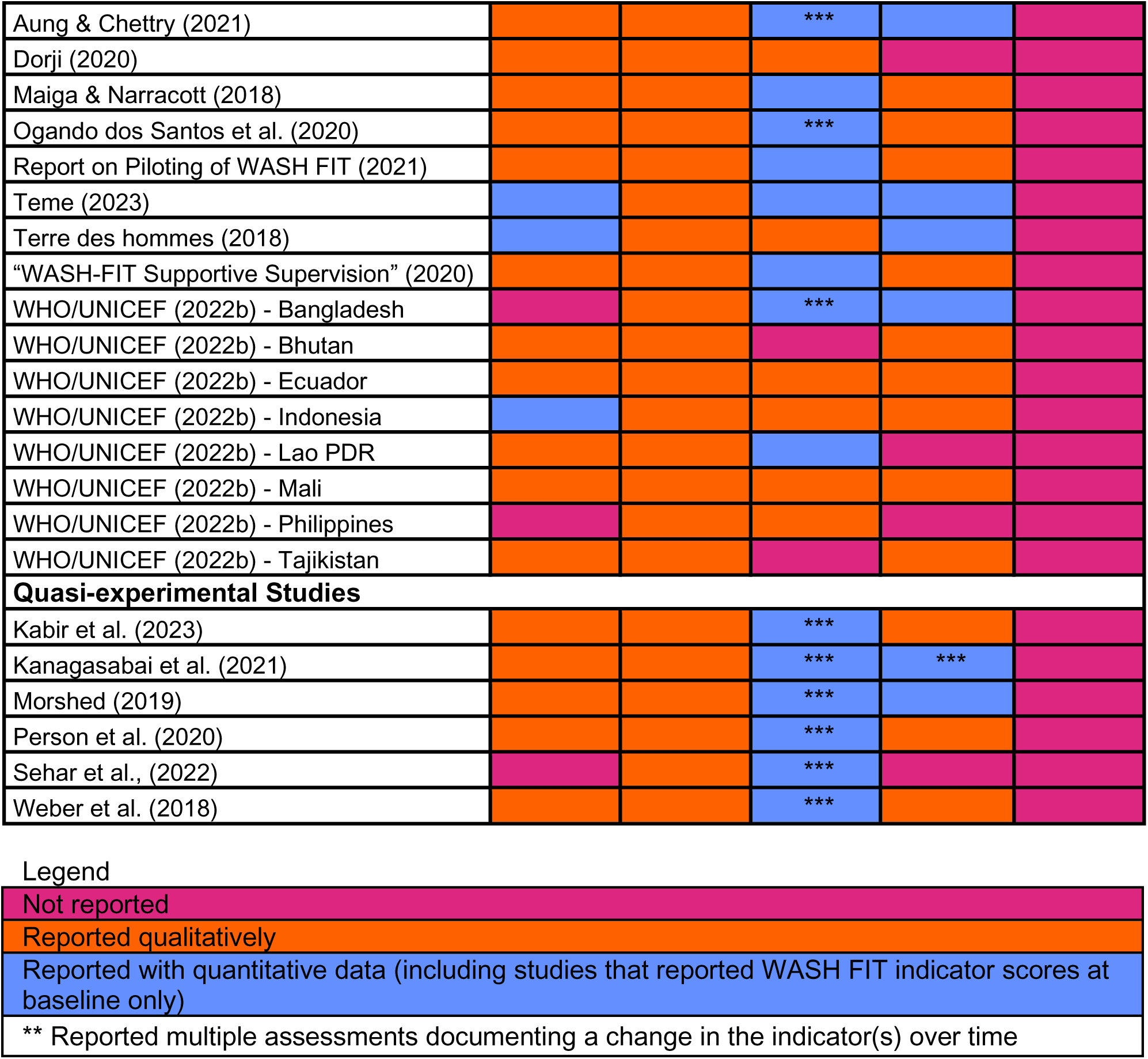
Reporting of inputs, activities, outputs, outcomes, and impacts among WASH FIT studies.

After reviewing similarities in our extracted data, we inductively developed the following input categories: financial support, government support, human resources, and technical assistance (Table 1). Financial support and human resources were reported both qualitatively and quantitatively, with five studies reporting specific funding amounts or numbers of staff, while others reported them qualitatively. Studies described government support and technical assistance qualitatively, though the level of detail varied. Some studies named specific government entities or international organizations, while others described receiving support from government entities or international organizations (Table 1).

**Table 1:** Categories of inputs for WASH FIT implementation.

| Category | Definition | Examples |
| --- | --- | --- |
| Financial Support | Financial or in-kind contributions of resources needed for WASH FIT implementation | Funds to establish the WASH FIT program (e.g., training of trainers, national-level orientation) (Kabir et al., 2023) (Terre des hommes, 2018) (Dorji, 2020)<br>Funds to make facility-level improvements (e.g., build infrastructure, purchase supplies) (Hirai et al., 2021) |
| Human Resources | Recruitment and/or training of qualified personnel to implement WASH FIT | Recruiting technicians and engineers for infrastructure improvements (Kanagasabai et al., 2021).<br>Training WASH FIT team (Ashinyo et al., 2021) (Saadeh et al., 2022) (Abdoulbaki Illio, 2023). |
| Technical Assistance | Subject-matter knowledge or expertise provided by an external entity to help address specific organizational needs | Supporting the Ministry of Health to co-develop a national infection prevention and control action plan (Weber et al., 2018)<br>Co-developing and leading a baseline assessment to target WASH FIT implementation with regional authorities (Morshed, 2019) |
| Endorsements and commitments | Endorsement, encouragement, or approval of project activities by regulators or champions | Endorsement of activities and verbal encouragement to WASH FIT teams from national-level ministry staff (Weber et al., 2018)<br>The town council approved the indicators for the WASH FIT assessment (Aung & Chettry, 2021). |

Like inputs, most studies reported activities qualitatively, describing the actions taken to implement WASH FIT. Studies did not directly tie activities to the WASH FIT cycle. Still, most reported activities could be mapped to the five steps in the cycle (i.e., establishing and training the WASH FIT team, assessing healthcare facilities, identifying and prioritizing areas for improvement, developing an improvement plan, and acting, monitoring, reviewing, adapting, and improving).

We also identified a commonly reported activity that was not included in the WASH FIT cycle but was suggested as an optional preparation in the WASH FIT guide. Ten studies reported adapting WASH FIT to local contexts or current needs. For example, the WASH FIT team in the Philippines consulted with stakeholders to harmonize WASH FIT indicators with national policies and guidelines (WHO/UNICEF, 2022b). Ogando dos Santos et al. (2020) translated and adapted WASH FIT material in Venezuela, and Teme (Teme, 2023) integrated 12 indicators from a COVID-19 scorecard into WASH FIT.

### 3.5. Outputs, outcomes, and impacts of WASH FIT

Most studies reported quantitative outputs related to WASH FIT implementation (n=23, 74%) (Figure 5). Outcomes were reported less frequently among all three study types. Eighty-three percent (n=5) of quasi-experimental studies, 76% (n=13) of case studies, and no cross-sectional studies reported outcomes. One case study reported qualitatively that staff perceived a reduction in “communicable diseases” after WASH FIT, but no study directly measured impacts. Based on observed similarities in our extracted data, we identified four output categories and seven outcome categories (Table 2).

**Table 2:** Categories of outputs and outcomes associated with WASH FIT implementation.

| Category | Examples |
| --- | --- |
| **Outputs** |  |
| Identification of WASH FIT gaps | Documenting the proportion of healthcare facilities meeting drinking water storage or bed spacing standards (Maina et al., 2019b).<br>Identifying healthcare facilities with lower levels of coverage for environmental health services compared to national and regional averages (Hirai et al., 2021) |
| Personnel trained or mentored on WASH FIT | Training or mentoring of staff on WASH FIT implementation (Report on Piloting of WASH FIT, 2021; WASH FIT supportive supervision, 2020)<br>Recruiting and training technicians to implement environmental health service improvements (Terre des hommes, 2018) |
| Infrastructure coverage and access | Improving existing infrastructure operations and maintenance (Aung & Chettry, 2021)<br>Constructing new or rehabilitating old infrastructure (Kanagasabai et al., 2021; Ogando dos Santos et al., 2021; Sehar et al., 2022; Weber et al., 2018) |
| **Outcomes** |  |
| Policy changes, including the decision to scale WASH FIT | Incorporating WASH FIT focal persons and improvement plans into national or regional standards for a minimal package of essential health services (Kabir et al., 2023)<br>Scaling up WASH FIT into the national strategy for the Ministry of Health (Weber et al., 2018)<br>Incorporating indicators for WASH FIT implementation into national health information monitoring systems (Teme, 2023) |
| Coordination, collaboration, and feedback | Enhancing collaboration between local government officials and healthcare facility-level administrators (Dorji, 2020; WHO/UNICEF, 2022b) |
| Funding for WASH FIT and environmental health services | Advocating at the national and regional levels for greater funding for WASH FIT and environmental health services (Weber et al., 2018; WHO/UNICEF, 2022b)<br>Incorporating WASH FIT into budgets at the healthcare facility level (Weber et al., 2018) |
| Knowledge, attitudes, and practices among healthcare staff | Improving staff motivation and satisfaction (Kabir et al., 2023; Weber et al., 2018)<br>Improving staff practices for infection prevention and control and waste management (Weber et al., 2018) |
| Knowledge, attitudes, and practices among patients/ community members | Improving staff practices for waste disposal in designated bins (Morshed, 2019)<br>Increasing community members' risk awareness and willingness to report issues to the WASH FIT team (Aung & Chettry, 2021) |
| Community engagement | Helping create infection prevention and control committees that included diverse participants and community members (Ogando dos Santos et al., 2021)<br>Engaging community members on the WASH FIT team (Morshed, 2019) |

We classified WASH FIT assessment indicators as output indicators because these indicators are designed to be assessed in Step 2 to inform improvement plans and then re-assessed in Step 5 to evaluate outputs and performance of the improvement plan. Most studies in our review reported conducting a facility assessment as Step 2 of the WASH FIT cycle. However, only six quasi-experimental studies and three case studies assessed and reported changes in WASH FIT indicator scores over time.

We identified an additional output indicator not included in the WASH FIT assessment tool: personnel trained on WASH FIT. Studies presented data about personnel trained on WASH FIT in varied ways, with some reporting the number of personnel trained while others only shared that an unspecified number received training.

Outcomes varied more widely than outputs and were less likely to be reported quantitatively. Ten studies reported that WASH FIT contributed to policy changes or informed governments’ decisions to scale up WASH FIT. Four studies reported that WASH FIT implementation helped improve collaboration with government entities. Seven studies reported that WASH FIT helped justify allocated budgets for WASH services. Twelve studies also noted improvements in occupational safety or knowledge, attitudes, or practices among healthcare staff, patients, or community members. Most of these changes were reported qualitatively through interviews or observations. Finally, four studies indicated that WASH FIT helped improve community engagement by creating new avenues and incentives for healthcare workers to engage with community members.

No studies rigorously reported impacts. One case study reported interview debriefing with program staff, stating that “two-thirds of the interviewees cited that there is a lower incidence of communicable diseases among the staff.” However, the methods did not specify how, if at all, communicable disease incidence was directly measured. Most studies referred to a connection between improved WASH conditions and various long-term impacts to justify the importance of WASH FIT. These included reduced healthcare-associated infections (Saadeh et al., 2022; Weber et al., 2018), antimicrobial resistance (Hirai et al., 2021; Sehar et al., 2021), infectious disease transmission (Ashinyo et al., 2021), and healthcare costs (Ashinyo et al., 2021; Morshed, 2019) as well as improved quality of care (Hirai et al., 2021; Sehar et al., 2022), health-seeking behaviors (Ashinyo et al., 2021; Hirai et al., 2021), and maternal and child health (Kanagasabai et al., 2021).

## 4. Discussion

This systematic review evaluated whether WASH FIT improves environmental health service-level outputs or health outcomes and impacts. Overall, we found insufficient evidence to answer this question. Only nine studies reported longitudinal data for outputs, all of which were quasi-experimental or case studies lacking a control group to attribute changes to WASH FIT implementation robustly. In nearly all studies, health outcomes were reported qualitatively or not at all, and health impacts were hypothesized but never directly measured. Our search yielded no experimental studies that robustly evaluated WASH FIT effectiveness on outputs, outcomes, or impacts.

Studies reporting longitudinal data on outputs did find beneficial changes, such as improvements in the proportion of healthcare facilities with access to basic water, sanitation, and waste management infrastructure (e.g., Kabir et al., 2023; Kanagasabai et al., 2021; Morshed, 2019; Weber et al., 2018). It is, therefore, plausible that WASH FIT improves service-level outputs. Continuous quality improvement tools have generally demonstrated favorable effects for other aspects of healthcare delivery (Knudsen et al., 2019; Kringos et al., 2015; Wells et al., 2018). However—given that many studies in our review were using WASH FIT as a one-time assessment tool rather than a continuous quality improvement tool—it is also plausible that initiating a WASH FIT program signals underlying political will and funding availability that would trigger improvements even in the absence of WASH FIT.

Studies in our review reflected positively on their experiences implementing WASH FIT and perceived changes in environmental conditions, which they attributed to WASH FIT. WASH FIT’s development and promotion by the WHO and UNICEF lend legitimacy and likely have contributed to its widespread popularity, uptake, and perceived effectiveness. However, we found that available evidence is too weak to conclude whether any observed improvements in environmental health services outputs or health outcomes and impacts can be attributed to WASH FIT.

Understanding the effectiveness of WASH FIT is critically important as it continues to grow in popularity and shape policies, priorities, and billions of dollars in investment. Given its influence, understanding if, how, and why it works should be prioritized for all stakeholders involved in funding, implementing, and promoting WASH FIT. Below, we discuss our findings in the context of three recommendations and the following steps to understand WASH FIT effectiveness better.

### 4.1. Disaggregate and report results of WASH FIT assessments

By design, implementing WASH FIT will yield quantitative data regarding output indicators for environmental health services (i.e., the results of the WASH FIT assessment tool). The WASH FIT guide recommends that healthcare facilities iterate an assessment cycle every 6-12 months. WASH FIT teams should review these data to guide improvement cycles and track the overall progress and effectiveness of the WASH FIT program (WHO/UNICEF, 2022a). As these data should be collected and managed in such a way as to allow routine review as part of the recommended WASH FIT process, including them in published studies should be straightforward. Yet, we identified several gaps in reporting.

Eighteen studies (58%) measured and reported quantitative data (i.e., WASH FIT indicator scores) for three or more WASH FIT domains or sub-domains. “Hand hygiene” and “management” were the domains least likely to be reported. Domains were not reported for several reasons. First, studies reported data from the WASH FIT first edition assessment tool for “hygiene” or “sanitation” without specifying which specific subdomains were measured for hand hygiene, environmental cleaning, sanitation, or waste management. This aggregation and ambiguity impeded our efforts to evaluate changes at the output level. However, the second edition of WASH FIT separates domains for hygiene, sanitation, cleaning, and waste management in the assessment tool, which should correct this issue. Second, studies indicated that they measured a domain but did not report data—or reported qualitatively that an output had changed but did not quantify specific metrics. In these cases, reporting checklists can help ensure that all essential information is reported, aiding the interpretation of individual studies and eventual meta-analyses (von Elm et al., 2007; Zwarenstein et al., 2008). Third, studies did not indicate whether they measured a specific domain and reported neither quantitative nor qualitative data. This may reflect a reporting deficiency or an adaptation to WASH FIT in which a particular domain was intentionally omitted from the assessment and improvement process.

We recommend that future studies disaggregate and report all WASH FIT assessment data. Documenting output changes is an essential preliminary step to understand whether and how downstream effects on health outcomes and impacts may occur. Furthermore, the effort required to track and report output-level changes should be minimal, as data collection and ongoing review are essential components of the WASH FIT cycle. Where multiple assessment rounds are conducted, studies should report the results of all assessments. Where programs intentionally modify or omit specific domains or indicators from their assessment and improvement plans, studies should explicitly describe the adaptation and rationale.

### 4.2. Identify inputs, activities, outputs, outcomes, and impacts

We used a logic model framework to identify inputs and activities required to implement WASH FIT and its resulting outputs, outcomes, and impacts. A logic model illustrates the core components of a program and outlines expected outputs, outcomes, and impacts that will be achieved when the specified inputs and activities are delivered (W.K. Kellogg Foundation, 2004). Logic models are an important tool for program planning and evaluation. They identify required inputs and activities to support process evaluations to understand if programs are being delivered as intended. They identify expected outputs, outcomes, and impacts, which can support impact evaluations to understand if programs are working as intended. Furthermore, logic models help understand a program’s theory of change (i.e., theory of how and why a program creates change, identifying specific pathways by which inputs and activities yield short-term outputs, intermediate outcomes, and long-term impacts) (Breuer et al., 2016; Savaya & Waysman, 2005; W.K. Kellogg Foundation, 2004).

Our review suggests that the underlying logic model and theory of change for WASH FIT are poorly documented or understood. We found selected examples of inputs, activities, outputs, outcomes, and impacts that were far from comprehensive. Furthermore, we did not identify studies that proposed links between specific inputs or activities and expected outcomes and impacts.

Other studies have hypothesized logic models. Within the WASH FIT guide, the WHO and UNICEF propose five input categories (political; financial/material; human; civil society engagement; intersectoral collaboration: energy and climate, health), activities for the five-step WASH FIT cycle, five outcome categories (improved staff morale and performance; increased care seeking; improved infection prevention and control and reduced antimicrobial resistance; less environmental pollution and more sustainable waste management; more efficient use of resources and lower healthcare costs), and three long-term impacts (dignified, safe pregnancy and reduced maternal and newborn mortality; healthier, more productive families and communities; improved outbreak response and resilience) (WHO/UNICEF, 2022a). Weber et al. (2019) proposed a conceptual evaluation framework for WASH FIT that closely follows the framework developed by WHO and UNICEF and additional outcomes for changes in finance and infrastructure operations and management. Our review identified similar categories, but overall, we found that the quality of evidence was too weak to corroborate the logic model. Furthermore, we identified many studies that did not complete all five steps of the WASH FIT cycle yet still attributed changes in outputs, outcomes, and impact to WASH FIT. This suggests that the underlying logic model and theory of change may differ from what has been proposed by the WHO, UNICEF, and others.

Understanding the logic model and underlying theory of change for WASH FIT is important for implementing and evaluating WASH FIT programs. It is also important to inform adaptations so that inputs and activities may be tailored to the context without undermining the core functions of the theory of change (Anderson et al., 2022; Kirk et al., 2020; Movsisyan et al., 2021). A well-defined logic model and theory of change can also enhance WASH FIT by guiding improvement plans based on local needs. For example, we hypothesize that hand hygiene and environmental cleaning are more likely to affect impacts related to surgical site infections and other healthcare-acquired infections compared to sanitation (Carboneau et al., 2010; Lederer et al., 2009; Pittet et al., 2000). Practitioners can develop improvement plans to target salient health issues if these pathways are understood.

We recommend research to define a logic model and theory of change for WASH FIT. Prior research on costing may help identify inputs and activities. Bottom-up costing studies on environmental health services in healthcare facilities have developed frameworks to identify essential inputs and activities (Anderson, Cronk, et al., 2021; Anderson, Wren Tracy, et al., 2021). However, these frameworks are not tailored to WASH FIT programs and will require verification and likely adaptation. Process evaluation or similar techniques (Saunders et al., 2016) to examine the implementation of WASH FIT programs will likely help develop and refine a logic model.

Research will also be needed to identify outputs, outcomes, and impacts. Output indicators are the best established, with 95 indicators already available in the WASH FIT second edition assessment tool (WHO/UNICEF, 2022a). Outcome and impact indicators will prove more challenging to evaluate. Prior systematic reviews have found insufficient evidence to assess the impacts of environmental conditions on healthcare-acquired infections (Watson et al., 2019) and patient satisfaction and healthcare-seeking (Bouzid et al., 2018). We expect systematic reviews for other outcomes and impacts to yield similar results, if not less conclusive.

We suggest that initial research should emphasize qualitative, mixed methods, and other formative work that identifies broad categories and specific types of outcomes and impacts rather than quantifying individual relationships. Health impact studies are needed but will be slow and costly. Qualitative and formative research can more rapidly and cost-effectively identify and map plausible benefits (e.g., Anderson et al., 2023). Once these broader frameworks have been established, systematic reviews can be used to compile evidence for specific impacts, generate meta-analyses, identify evidence gaps, and develop a more precise research agenda.

### 4.3. Conduct experimental studies and exploit natural experiments

Experimental studies will be necessary to rigorously evaluate WASH FIT’s effectiveness in improving health outcomes and impacts. These studies will be costly. However, given the influence and investment in WASH FIT by countries—both countries adding line items for WASH FIT into their national health systems budgets and bilateral donors—we argue that they are important to ensure these investments are effective and identify improvement opportunities.

In parallel to experimental studies, there are opportunities to exploit natural experiments among countries’ WASH FIT programs using national health management information systems. Reducing healthcare-acquired infections is routinely cited as a benefit of WASH FIT (Ashinyo et al., 2021; Hirai et al., 2021; Saadeh et al., 2022; Sehar et al., 2021; Weber et al., 2018).

Measuring healthcare-acquired infections is expensive and requires large sample sizes for experimental trials (Blanco et al., 2019). However, many countries monitor healthcare-acquired infections as part of their national health management information systems. In countries implementing WASH FIT in a subset of regions or healthcare facilities within a region, matched controls could be identified among non-program areas to evaluate impact using existing data more rigorously. Other health outcomes and impacts could be assessed through proxies such as changes in patient volume or facility revenue to approximate changes in care seeking (Lopez et al., 2020).

We recommend that country governments, implementing partners, and any external funders incorporate more rigorous evaluation plans into WASH FIT programs. One possible solution to support countries and implementing partners is to incorporate evaluation guidelines into the WASH FIT implementation guide or create a companion guide as part of the suite of available WASH FIT materials. WHO and UNICEF have created supplemental materials, including a training manual and technical fact sheets (WHO/UNICEF, 2022c). An evaluation guide could supplement these materials. We recommend that an evaluation guide encourage WASH FIT teams to document inputs and activities needed to implement WASH FIT and report on output, outcome, and impact-level indicators to help strengthen causal pathways and evaluate the WASH FIT’s long-term effectiveness.

### 4.4. Limitations

We limited our search to two peer-reviewed databases and one grey literature database. Washinhcf.org is the most comprehensive grey literature database for environmental health services in healthcare facilities, but it does not actively solicit studies. Our review captured available published studies, but there are likely many unpublished reports. Dedicated efforts to make those reports publicly available would strengthen the evidence base.

We recognize that definitions to classify outputs, outcomes, and impacts vary. Our review adapted definitions from the Kellogg Foundation (2004). We classified extracted data accordingly, but arguments could be made to classify some long-term outcomes (e.g., policy changes) as impacts. Vague descriptions hindered our efforts to extract data into the logic model framework in the literature. For example, studies that reported that a specific organization or government agency gave “support” were classified as inputs. However, this could be classified as an output if the “support” indicates knowledge, attitudes, or policy-level action changes after WASH FIT implementation.

## 5. Conclusions

This systematic review sought to evaluate WASH FIT’s effectiveness at improving environmental health service-level outputs and health outcomes and impacts. Lack of quantitative data (particularly for outcomes and impacts), incomplete and inconsistent reporting, and weak study designs posed challenges. Most studies reported outputs in terms of indicators measured as part of the WASH FIT assessment tool. Still, many of these assessments were done to initiate the WASH FIT cycle and were not followed up with assessments after implementing WASH FIT improvement plans. We found no studies used a control group that would demonstrate change over time attributable to WASH FIT. WASH FIT may plausibly improve service-level outputs. However, WASH FIT programs may reflect commitments and investments in environmental health services, yielding similar outputs even without WASH FIT. In either case, we cannot determine whether these service-level outputs improve health outcomes and impacts.

Understanding whether and how WASH FIT achieves impact is important and will require future research. This evidence is important to ensure that funding invested for WASH FIT implementation is used cost-effectively and that opportunities to adapt and refine WASH FIT are fully realized as it continues to grow in popularity and influence. As a first step, we encourage more transparent reporting of all output indicators captured in WASH FIT assessments, particularly for follow-up and long-term monitoring after initial improvement plans are implemented. In the long-term, we recommend experimental studies and exploiting natural experiments where staggered implementation or sub-national programs operating only in selected regions offer the opportunity to compare the WASH FIT program versus non-program areas using data within health management information systems on healthcare-acquired infections and other health impacts tracked within health management information systems.

## Funding

This work was funded in part by a grant from the Wallace Genetic Foundation to the Water Institute at UNC. Darcy Anderson is supported by a grant from the National Institute of Environmental Health Sciences (NIEHS) (T32ES007018).

## Declaration of competing interests

None.

## Data Availability

All data produced in the present study are available upon reasonable request to the authors

## Acknowledgments

This research was conducted by researchers at the Water Institute at UNC, and we thank Kaida Liang and other Water Institute staff for their support.

## Author contributions

**Hannah Lineberger:** Writing – original draft, Visualization, Analysis. **Ryan Cronk:** Writing – review and editing, Supervision, Conceptualization. **Sena Kpodzro**: Analysis, Writing - review and editing. **Aaron Salzberg**: Funding acquisition, Writing - review and editing. **Darcy Anderson:** Writing – review and editing, Supervision, Conceptualization.

## Notes

### Competing Interest Statement

The authors have declared no competing interest.

